# Arboviruses in Kenya: A Systematic Review and Meta-analysis of Prevalence

**DOI:** 10.1101/2024.10.17.24315511

**Authors:** Lynn J Kirwa, Hussein M. Abkallo, Richard Nyamota, Enock Kiprono, Dishon Muloi, James Akoko, Jennifer S. Lord, Bernard Bett

**Author notes:** Corresponding author: Lynn J Kirwa.

## Abstract

Arboviruses cause >700,000 human deaths annually, with Rift Valley fever (RFV), yellow fever (YF), chikungunya, and dengue outbreaks posing major public health and economic challenges in East Africa. Yet, no comprehensive studies have consolidated Kenya’s historical arboviral data to support risk assessment and inform control strategies. We registered this review in PROSPERO (CRD42023407963) and searched Web of Science, PubMed, and Global Health databases for observational articles reporting prevalence from the three main arboviral families from inception until 15^th^ March 2023. We pooled the IgG prevalence of arboviruses using a random-effects meta-analysis with a generalised linear mixed-effects model. Heterogeneity was assessed and quantified using Cochran’s Q and *I^2^* statistics and 95% prediction intervals estimated. We included 65 articles (246 datapoints; 14 arboviruses) in our analysis. The pooled IgG prevalence of RVF was 16% (95% CI: 11–24%; I²=70%) in wildlife, 10% (95% CI: 8–13%; I²=90%) in livestock, and 7% (95% CI: 4–11%; I²=98%) in humans, with consistently high rates observed in Garissa and Tana River counties. Among Aedes-borne viruses, chikungunya showed the highest prevalence (10%; 95% CI: 4–24%; I²=99%), followed by dengue (6%; 95% CI: 3–11%; I²=98%) and YF (5%; 95% CI: 2–11%; I²=97%), with the highest prevalence in Busia and Kwale. West Nile virus prevalence in humans was also estimated at 9% (95% CI: 5–14%; I²=93%). Overall, the Coast, Western, and Rift Valley regions were the most affected. Multiple arboviruses have historically circulated Kenya, and with the increasing pressures of climate change, urbanization, and global connectivity, the risk of outbreaks, particularly from Aedes-borne viruses, is escalating. Proactive, sustained surveillance as well as integrated public health strategies through a One Health lens are needed to mitigate these threats and protect vulnerable populations.

**Author Summary:** Infections like Rift Valley fever, dengue, chikungunya, yellow fever, and West Nile virus are major neglected tropical diseases that pose significant public health threats both in Kenya and globally. Given the conducive conditions in Kenya and the absence of a comprehensive routine surveillance system, many areas remain under-monitored, increasing the risk of undetected transmission and delayed responses, which leaves vulnerable populations at greater risk. It is therefore imperative to conduct a thorough assessment of baseline prevalence for these diseases through a systematic review and meta-analysis that consolidates and analyses existing evidence for improved public health planning. Our study provides a detailed review of historical IgG prevalence data for 14 of these diseases in Kenya, illustrating their long-standing circulation in human, livestock, and wildlife populations, as well as the heightened risk of outbreaks driven by climate change and urbanization. We emphasize the urgent need for ongoing surveillance and integrated public health strategies tailored to Kenya’s unique context, such as those supported by the GAI initiative, to effectively protect vulnerable populations from future outbreaks, nationally and regionally.

## Introduction

Vector-borne diseases constitute *approximately* 17% of the estimated global burden of infectious diseases and cause >700,000 deaths annually (1,2). Arthropod-borne viruses (arboviruses) contribute substantially to this burden, with dengue virus (DENV) alone responsible for >6.5 million human infections and >7300 deaths in 2023 (3). With severe associated complications like dengue shock syndrome and long-term chikungunya disability, potential epidemiological shifts due to changes in climate and vector distribution, in addition to urbanisation, there is an urgent need for increased arboviral research and management (2,4–7). In response, WHO launched the Global Arbovirus Initiative (GAI) to build a coalition of countries and strategic multisectoral partners, to improve surveillance, prevention, and control through an integrated and coordinated approach for maximum impact in high-burden and at-risk areas (8,9). The GAI‘s first pillar, *’monitor risk and anticipate*’ emphasizes developing a robust surveillance system that integrates current and historical data to establish baselines, and mapping risks to identify vulnerable areas. Studies on IgG exposure support these priority actions by contributing to establishing local historical baselines and improving data quality to identify areas at risk, allocate resources efficiently, and formulate innovative targeted mitigation strategies (9).

In Kenya, like many tropical countries, human and animal populations are vulnerable to arboviruses owing to the optimal climate that supports a wide range of vector species able to transmit arboviruses (10). Several arboviruses, both well-established and emerging, including DENV, chikungunya (CHIKV), West Nile (WNV), Rift Valley fever (RVFV), Ngari (NV) and yellow fever (YFV) have been documented in the country (10–13). Furthermore, Kenya’s proximity to countries reporting recurring arboviral outbreaks like Uganda, Sudan, Tanzania, and Somalia may exacerbate the risk of arboviral importation into the country through transboundary movements (14,15)Despite these irrefutable risks, Kenya lacks a comprehensive routine surveillance system to actively monitor arboviral activity across the country for timely intervention (15,16). Existing surveillance efforts in Kenya are reactive, intensifying once outbreaks are underway or through interepidemic investigations, often focusing on high-profile arboviruses and identified hotspots (17). Consequently, large parts of the country may be under-monitored, increasing the potential for undetected transmission and delayed responses, leaving vulnerable populations at greater risk. Considering the healthcare burden they and other vector-borne diseases exert, alongside their impoverishing socioeconomic ramifications (18,19), the urgency for a thorough assessment of their baseline prevalences in the country per WHO-GAIV goals cannot be overemphasised. Here, we report the findings of a systematic review and meta-analysis of IgG seroprevalence in humans, livestock, and wildlife, consolidating and analysing existing evidence on arboviral exposure across Kenya. Given Kenya’s position as a regional trading, transportation and healthcare hub, our findings will contribute to national research and control efforts in addition to regional strategies for arboviral disease mitigation.

## Methods

Our systematic review and meta-analysis protocol was registered in advance in the PROSPERO International Prospective Register of Systematic Reviews (CRD42023407963) and was reported according to the Preferred Reporting Items for Systematic Reviews and Meta-analyses guidelines (S1 File) (20).

### Systematic search of the literature

We conducted a systematic search in PubMed (since 1946), Web of Science (since 1900), and Global Health (since 1973) databases from their inceptions to 15^th^ March 2023. Our search syntax (S2 File) was based on keywords from the Medical Subject Headings (MeSH) database and was designed to capture all published studies reporting prevalence data for arboviral families, including *Togaviridae* (CHIKV, ONNV, Sindbis virus (SINV)), *Flaviviridae* (DENV, WNV, YFV, ZIKV, TBEV), *Bunyaviridae* (Orthobunyavirus, NV, RVFV, CCHFV), and other related viruses in Kenya. All selected articles were collated and uploaded into Rayyan QCRI (21) and filtered for duplicate records.

We considered cross-sectional studies, both community- and hospital-based, before, during and after reported arboviral outbreaks in Kenya. We included studies that reported prevalence of arboviruses in human, livestock and/ or wildlife populations based on serological detection of immunoglobin G or M (IgG, IgM), viral isolation or reverse transcription–polymerase chain reaction (RT-PCR). We excluded all studies conducted outside Kenya, and any comments, conference notes, reviews and meta-analyses, case reports, and non-peer-reviewed manuscripts (21) Two (L.J.K., R.N.) reviewers independently and blindly screened the remaining articles in two stages: primary screening of the titles and abstracts, and secondary screening of full-text articles, recording reasons for exclusions (21). Disagreements were resolved by discussing with two additional independent reviewers (H.A., J.L.).

### Evaluation of the quality of studies

Two reviewers (L.J.K., R.N.) independently assessed each included study’s quality using the Joanna Briggs Institute System for the Unified Management, Assessment and Review of Information (JBI SUMARI) critical appraisal tool (S3 File) (22). Specifically, this appraisal was done against nine criteria encompassing elements related to sampling, population representativeness and description, sample size determination, diagnostics coverage and availability, statistical analysis, and addressing low response rates (23). Disagreements were resolved by discussing with two additional independent reviewers (H.A., J.A.). Study quality was scored as high (7-10), moderate (4-6), or low (0-3), with studies rated as high and moderate considered suitable for inclusion in the meta-analysis (24–26).

### Data extraction

Three reviewers (L.J.K., R.N., E.K.) extracted data from included studies, to a single predefined summary Excel sheet (S4 File). These data included the first author, publication year, study design, sampling method (probabilistic, non-probabilistic), sample size, arbovirus, study setting (rural, urban, peri-urban), study period, transmission period (during or between outbreaks), geographical location (county, region), population, human participant recruitment (community, health facilities), human clinical status (apparently health, febrile illness), detection assay, confirmatory test if any, and proportion of positives per total numbers tested. In publications reporting seroprevalence studies from more than one arbovirus, population, and/ or geographic locations (counties or regions), each was considered as a separate study/ row of data.

### Meta-analysis of prevalence

For each study where IgG data were available, we calculated IgG prevalence estimates and their corresponding 95% confidence intervals using the exact binomial method (28). We then conducted a meta-analysis of prevalence for each arbovirus using R statistical software (version 4.2.0), using the ‘meta’ package. We estimated the pooled prevalence for each arbovirus and host population using a random intercept logistic regression model with a logit link function. The magnitude of heterogeneity between studies was assessed using Cochran’s Q and quantified using the *I*^2^ statistic; with an *I*^2^ of more than 75% indicating substantial heterogeneity in the studies (27). We estimated 95% prediction intervals to capture the expected range of true proportions, and used forest plots to visualise the results (28). The distribution of arbovirus IgG prevalence across different geographical locations was mapped using QGIS software (version 2.18.26) (31).

## Results

### Study selection

We retrieved 3085 publications and after removing 1055 duplicates, 2030 were screened by title and abstract. From these, 97 full texts were screened, resulting in 65 articles that met our inclusion criteria (Fig. 1). Thirty-seven of these articles included multiple seroprevalence studies for more than one arbovirus, population, and/or geographic location (counties or regions) and were thus considered separately for analysis, resulting in 246 datapoints. Most studies (85%, 209/246) were of high quality, with 15% (37/246) rated as moderate quality (S5 File).

**Fig. 1.** Study selection process based on the PRISMA flowchart. Abbreviations: n = number of studies.

### Summary of seroprevalence studies

Of the studies, 40.7% (100) reported IgG seroprevalence for RVFV, followed by DENV (15%, 37), WNV (8.9%, 22), CHIKV (8.9%, 22), YFV (6.1%, 15), ZIKV (4.5%, 11), TBEV (4.1%, 10), CCHFV (4.1%, 10), ONNV (2.8%, 7), Sindbis (SINDV) (1.2%, 3), Banzi (BANZV) (1.2%, 3), Bunyamwera (BUNYV) (1.2%, 3247), NV (0.8%, 2), and Orthobunyaviruses (0.4%, 1). Of these studies, 74.8% (n=184) focused on human populations, 14.2% (n=35) included livestock sampling and 11% (27) included wildlife.

Among 184 human population studies, 64.7% (n=119) sampled apparent healthy individuals, while 31.5% (58) sampled those presenting in health facilities with febrile illnesses that were confirmed as arboviruses: DENV (17), WNV (11), TBEV (7), CCHFV (5), CHIKV (5), RVFV (4), YFV (4), ZIKV (4), and Orthobunyaviruses (1). Seven (3.8%) studies were carried out in both settings. Most of the studies were cross-sectional (97.2%, 239), with the remainder being cohort (1.6%, 4) and longitudinal (1.2%, 3). Additionally, 64.2% (158/246) carried out between outbreaks with human participants recruited from either the community (119/184, 64.7%) or health facilities (65/184, 35.3%). All eight regions in Kenya were represented by at least one study (Fig. 2). Coast and Rift Valley regions region had the highest number of studies while Nairobi and Central regions had the least.

**Fig. 2.** Number of publications reporting seroprevalence estimates by geographical region: RVFV (*A*); DENV (*B*); YFV (*C*); CHIKV (*D*). Source: The base map was retrieved from https://data.humdata.org and modified with QGIS software version 3.36.1-Maidenhead.

Of the 246 studies, IgG antibodies were the most tested (41.1%, 101/246), followed by combinations of IgG and IgM (27.6%, 68/246), unspecified antibodies (15.4%, 38/246), IgM and viral RNA (4.9%, 12/246), IgG/IgM (4.5%, 11/246), IgG/IgM and viral RNA (1.2%, 3/246), IgA, IgM, and IgG (1.6%, 4/246), IgM (1.6%, 4/246), and arboviral RNA (1.6%, 4/246). Most arboviral diagnostic assays across the studies were serological such as ELISA (50.8%, 125/246), hemagglutination inhibition (HI; 11.4%, 28/246), plaque reduction neutralization test (PRNT; 6.9%, 17/246), and indirect immunofluorescence assay (IIFA; 1.2%, 3/246) while some used molecular tests including RT-PCR (2.4%, 6/246) and sequencing (5.7%, 14/246). A few studies combined both methods, such as ELISA and PRNT (4.5%, 11/246), ELISA, PRNT, and IIFA (0.8%, 2/246), RT-PCR and sequencing (0.4%, 1/246), ELISA and RT-PCR (9.8%, 24/246), and ELISA, RT-PCR, and sequencing (4.9%, 12/246). Among these, 22.8% (56/246) carried out further confirmatory testing: PRNT (42/246; 17.1%), virus neutralization test (VNT; 10/246; 4.1%), FRNT (3/246; 1.2%) and ELISA (1/246; 0.4%) (S6 File).

The studies included in our analysis were undertaken between 1966 and 2021, with the earliest eligible study published in 1970, followed by studies in 1982, 1983, and 1992, totalling to 28 studies. These studies focused on RVFV, CHIKV, CCHFV, SINV, BUNV, ZIKV, YFV, BANV and WNV. Post-2000, the next eligible study was published in 2002 (n=1) and then 2007 (n=11), all focusing on RVFV. There was an increase in published studies from 2007 onwards, peaking in 2010 (n=28), 2015 (n=45) and 2020 (n=37), which coincided with RVFV and DENV outbreaks (Fig. 3). Overall, RVFV was the most consistently studied arbovirus during this entire period, with publications in all identified years except 1970 and 2012. Other arboviruses most frequently studied were CHIKV, DENV, WNV, and YFV (S6 File).

**Fig. 3.** Number of publications reporting seroprevalence estimates in humans, livestock and/ or wildlife, for each arbovirus by publication year.

### Reported IgG prevalence of arboviruses in Kenya RVFV

We calculated a pooled IgG seroprevalence of 7% (95% CI: 4 - 11%; I^2^: 98%, number of studies: 33) and a prediction interval of 0- 62%, reflecting substantial heterogeneity among the studies in humans. Prevalence ranged from zero in Uasin Gishu (Rift Valley) in 1996 and Kilifi (Coast) in 2004 to a 40% in Nakuru (Rift Valley) in 1989 (Fig. 4A). In livestock, twenty-seven studies reported a pooled prevalence of 10% (95% CI: 8 - 13%; I²: 90%) (Fig. 4B). The prediction interval ranged from 3% to 30%, with the highest prevalence observed in Garissa (North Eastern) in 2015 at 30% and the lowest in Samburu (Rift Valley) and Meru (Eastern), both at 2% in 2006. We observed an IgG seroprevalence of 16% (95% CI: 11 - 24%; I^2^: 70%, n=15) in wildlife, with a prevalence range from 5% in Taita Taveta and Tana River (both in Coast) in 2007 and 2008, respectively, to 57% in Garissa (North Eastern) in 2007, and a prediction interval of 4% to 51% (Fig. 4C). The most frequently sampled species were buffaloes, warthogs, oryx, waterbuck and impala.

**Fig. 4.** 2Forest plots showing meta-analysis of prevalence for Rift Valley fever virus. A: humans; B: livestock; and C: wildlife. The first column lists the author and year of study, followed by the area (county/region) in Kenya. Error bars represent the pooled proportions/prevalence of IgG antibodies with their 95% CIs on the log scale for each study. Diamonds indicate the combined point estimate. I² statistics, τ², and Q-test p-values are reported.

### *Aedes*-borne viruses

Of 21 studies focusing on humans, the pooled IgG seroprevalence for DENV was 6% (95% CI: 3–11%; I² = 98%), with a prevalence range from zero in Busia (Western) in 2004 to 67% in Kwale (Coast) in 2000 and 60% in Coastal counties in 2007, and a prediction interval of zero to 67% (Fig. 5A). In wildlife, specifically *Papio* (baboon) species, a pooled prevalence of 27% (95% CI: 12-49; I^2^ = 81%) was estimated from two studies, with a prediction interval of 8-61%, the highest prevalence being in Kwale (Coast) at 47% and the lowest in Kakamega (Western) at 15%, both reported in 2014.

**Fig. 5.** Forest plots showing meta-analysis of prevalence for Aedes-borne viruses. A: Dengue virus; B: Chikungunya virus; and C: Yellow fever virus. The first column lists the author and year of study, followed by the area (county/region) in Kenya. Error bars represent the pooled proportions/prevalence of IgG antibodies with their 95% CIs on the log scale for each study. Diamonds indicate the combined point estimate. I² statistics, τ², and Q-test p-values are reported.

We reviewed 15 studies reporting CHIKV IgG seroprevalence in humans, showing a pooled prevalence of 10% (95% CI: 4-24%; I² = 15), and a prediction interval of 4-24%. Prevalence ranged from 72% in Lamu (Coast) in 2004 and 67% in Busia (Western) in 2010, to a low of 1% in Nairobi (2008), Central (2007), and Kwale (Coast) in 2009 (Fig. 5B). There was an overall IgG prevalence of 5% (95% CI: 2-11%; I^2^= 97%, n = 9) for YFV in human populations. The prediction interval was 4-24%, with a prevalence range from a high of 42% in Kwale (Coast) in 2000 to a low of 1% in Kilifi (Coast) and Samburu (Rift Valley), both reported in 2004 (Fig. 5C). The pooled prevalence of IgG ZIKV in humans, from four studies was 2% (95% CI: 1-3%; I^2^=0%; prediction interval: 0-5%). Reported prevalences ranged from 3% in Kisumu (Nyanza) in 2011 to 1% in Nairobi and Kisumu (Nyanza) in 2009.

### Other mosquito-borne viruses

Pooling data from seventeen studies on WNV IgG, we report a pooled prevalence rate of 9% (95% CI: 5-14%; I²=93%), with a prediction interval of 1-51%. The highest prevalence rates were in Tana River in 2009 (34%) and Kwale in 2000 (29%), both in Coast and the lowest in Samburu (Rift Valley) (1%) and Kilifi (Coast) (2%), both reported in 2020. For ONNV, an overall prevalence of 3% (95% CI: 1 - 9%; I^2^= 76%; n=7) was observed, with a prediction interval of 0- 40%, the highest prevalence being in Lamu (Coast) at 20% in 2008 and the lowest in Nairobi at 1% in 2008. From the only two studies in Rift Valley region in 2022 reporting IgG Ngari in livestock, a pooled prevalence of 38% (95% CI: 31- 46%) with a moderate heterogeneity (I^2^= 30%) was observed. The prediction interval was 31-46%, with individual prevalence rates of 31% in Kajiado and 24% in Baringo.

### Tick-borne viruses

An overall human IgG TBEV prevalence of 7% (95% CI: 5 - 10%; I^2^=79%) was estimated from ten studies, with a prediction interval of 2-20%, indicating moderate heterogeneity. Prevalence rates ranged from 16% in Kwale (Coast) and 11% in Baringo (Rift Valley) counties in 2000 and 2020, respectively, to 3% in Nandi (Rift Valley) in 2000. For CCHFV, pooled estimates of IgG seroprevalence in humans were 15% (95% CI: 6–33%; I^2^=97%; n=5), with a prediction interval of zero to 90%. The highest prevalence was reported in Baringo (Rift Valley) in 2009 at 35%, while the lowest was 3% in a nationwide study in 2020. In livestock and wildlife, we report pooled CCHFV prevalence rates of 28% (95% CI: 21–36%; I^2^=0%; n=1) in Laikipia and Narok counties in Rift Valley, and 75% (95% CI: 69–81%; I^2^=0%; n=1) in Meru (Eastern) and Laikipia, Narok, Nakuru counties in Rift Valley, all reported in 2007.

## Discussion

Our systematic review and meta-analysis, provide insights into the historical occurrence of 14 arboviruses in humans and animals. We focus on RVFV, DENV, CHIKV, YFV, and WNV due to their substantial prevalence and prioritization in key global initiatives like the WHO Blueprint for Prioritized Diseases.

Our findings indicate widespread RVFV seropositivity across humans, livestock and wildlife that is potentially associated with presence of competent vectors, including *Aedes*, *Culex*, *Mansonia*, and *Anopheles* spp. (10,30), alongside livestock movements related to intensive pastoralism, commercial and sociocultural practices (31–34). The higher seroprevalences in livestock and wildlife than humans are as expected due to more frequent livestock-vector-wildlife transmission, with less common spillover to humans, but still of concern (35). A One Health approach remains vital for monitoring these dynamics and mitigating accordingly. We note that the majority of studies have been concentrated in North Eastern (Garissa, Wajir), Coast (Tana River, Kilifi, Kwale, Taita Taveta), and Rift Valley (Nakuru, Baringo). This focus likely reflects targeted research in “at-risk” areas rather than limited risk elsewhere, thus a need to expand surveillance to under-studied areas to fully capture the national risk (36) For DENV and CHIKV in humans, we report overlapping geographic distributions in Nairobi, Nyanza, Western, and coastal counties (10,30), and indicating risks of cocirculation/ coinfection, particularly in areas with abundant *Aedes aegypti* mosquitoes. High frequency of outdoor livelihood activities, including pastoralism, farming, and fishing, across much of the country could further increase exposure risk to the day-feeding vector, supporting our findings (37,38). Wildlife may play an important role in DENV transmission in forested areas such as Kwale and Kakamega, either through spillback from humans to nearby *Papio* species (16,38–40) or via enzootic transmission driven by sylvatic *Aedes aegypti formosus* (41–43). YFV’s relatively low seroprevalence herein was unexpected, especially given Kenya’s designation by WHO as a high-risk country and routine mass vaccination campaigns albeit only in high-risk counties (44,45). With Kenya’s human YFV vaccination coverage estimated at just 7%, far below WHO-recommended 80% for herd immunity (10,45,46), undetected YFV circulation in susceptible populations alongside competent *Aedes* spp. (10,30) may explain our findings. However, since the most recent data in our study is from 2017, and dynamics may have shifted, especially after the 2022 outbreak, reassessing YFV prevalence is key in light of current trends. Finally, with no documented outbreaks, WNV substantial seroprevalence in humans suggests sporadic transmission likely linked to migratory birds at convergence zones or local mosquito vectors, including *Culex, Mansonia* and *Aedes* spp (10,30,47,48).

Our findings illustrate a persistent, widespread risk of arboviral transmission in Kenya, one that we expect to escalate due to evolving environmental and socioeconomic conditions, including climate change, globalization, urbanization, and rapid population growth (7,49–52). With respect to climate change, heavy rainfall and flooding events influence RVFV (53–57), warmer temperatures and altered precipitation patterns for *Ae. aegypti*-mediated DENV, CHIKV, and YFV (58–62), and prolonged dry spans and droughts necessitating water storage in containers for DENV and CHIKV (62–64). As climate change intensifies these weather patterns, permissive vectors may expand their geographical range and transmission seasons, potentially exacerbating arboviral threats (49,65). Under the latest IPCC projections, Kenya and much of East Africa is expected to experience mean annual temperature increases of up to 2.1°C (57) with intensified heavy rainfall events (57,66–69) and sustained or reduced drought frequency (70–73). Consequently, extensive outbreaks in established hotspots could worsen, and previously unaffected or less affected regions in Kenya may emerge as new hotspots. Mordecai et al. suggest that climate-driven shifts could create complex changes in the incidence of arboviruses and malaria, with warmer temperatures potentially favoring one over the other depending on the region (51). In Kenya, this could worsen the dual burden of arboviruses and malaria, additionally straining the health system and complicating public health efforts. Rapid, often unplanned urbanization, characterized by mushrooming slums, overcrowding and poor sanitation, provides conducive breeding conditions for *Ae. aegypti* and *Culex pipiens*, which could facilitate the rapid spread of DENV, CHIKV, ZIKV and WNV (7,74–76). This is reflected in our findings of considerable prevalence of Aedes-borne viruses in urban centers like Nairobi, Mombasa, Kisumu, Busia, and Malindi. This transmission risk may be further compounded by the urban heat island effect in Nairobi, and other cities that could enhance mosquito survival and reproduction, while the urban sprawl could potentially hamper vector control measures like larval source management (77–79). As Kenya’s cities and towns continue to expand and with more than 50% of Kenyans projected to live in urban centres by 2030 (80,81), the threat of urban arbovirus outbreaks in the country is significant. Further, *Ae. albopictus*, a highly adaptable species, was recorded in Kenya’s neighbors nearly a decade ago (7,49–52) and could already be in Kenya undetected (82), raising concerns about its potential to facilitate the spread of arboviruses in both urban and rural settings, further complicating control efforts.

Beyond expansion of urban and peri-urban centres, deforestation, intensive pastoralism, land-use change and encroachment of human settlements into previously uninhabited areas could also increase arbovirus risk by disrupting natural ecosystems and increasing human-sylvatic vector interactions (83,84). Additionally, globalization—through increased international travel, tourism and trade—compounds these risks by facilitating the movement of infected people and livestock across borders, potentially introducing new strains of arboviruses into Kenya and triggering outbreaks in previously unaffected regions (85–88). Our findings support this, with substantial representation of Aedes-borne viruses in the trade and tourism-intensive coastal counties of Mombasa, Kwale, Kilifi, and Lamu.This is particularly concerning for viruses like DENV and CHIKV, which have a propensity for rapid spread in new environments (89–91), and is especially relevant for coastal counties like Mombasa, where high volumes of international goods importation and tourism could also introduce invasive mosquito species like *Ae. albopictus*.

Mitigating these projected surging arboviral threats in Kenya requires aligning national efforts with global frameworks like the GAI, Global Vector Control Response (GVCR), and WHO Regional Framework for monitoring invasive species (92–94)Kenya’s surveillance infrastructure, particularly the Integrated Disease Surveillance and Response (IDSR) system, has traditionally relied on passive surveillance methods with marked successes in detecting outbreaks like cholera and malaria (95,96). It is, however, often limited by underreporting and delays, limiting its capacity to fully monitor arboviral activity across the country (97) Further, deficiencies in diagnostic capacities, data integration across human, animal, vector and environmental health sectors, and insufficient personnel training have emerged as limiting factors in Kenya’s capacity for arboviral surveillance and control (98). To bridge these gaps, Kenya needs to incorporate more active surveillance methods within a One Health framework including expanding entomological surveillance, streamlining reporting mechanisms, and leveraging geographic information systems and remote sensing technologies that could improve Kenya’s ability to monitor vector populations and predict outbreak hotspots before they escalate (92,99) More focus on arbovirus co-infections and their clinical outcomes is needed in Kenya, as most included studies examined single viruses at a time, despite many vectors like *Aedes* and *Culex* spp. often harbouring multiple viruses simultaneously (100). Future research should also prioritize longitudinal studies across diverse ecological zones to comprehensively assess arbovirus circulation and transmission dynamics across the country. Further, diagnostic challenges such as overlapping clinical symptoms, cross-reactivity, and persistence of antibodies among many arboviruses limit the ability of laboratories in the region to generate accurate exposure data. Incorporating more specific case definitions, along with expanding the diagnostic capabilities — accurate point-of-care diagnostics especially in remote, resource-limited settings — is key for timely outbreak detection and management (51,101).

With no vaccines available for most arboviruses, integrated vector management (IVM) remains the most viable approach for outbreak prevention and response (89). In Kenya, existing vector control efforts, including larval source management (LSM), insecticide-treated nets (ITNs), and indoor residual spraying (IRS), were primarily developed to combat malaria and have successfully reduced its burden (102,103). Yet, this focus has left arboviruses largely neglected. To correct this imbalance while maximizing resource efficiency, Kenya can leverage these existing infrastructure and resources to control arboviruses, adopting them to specific vector needs (89). For example, while LSM and larvicides may apply across *Anopheles* (malaria) and some *Aedes* species, ITNs may be less effective due to vector biology and behavior differences (89,104)Further, given Kenya’s dual burden, these arbovirus interventions should complement and not detract from malaria control achievable, for instance, by training malaria control experts in arbovirus management and securing funding agreements to prevent resource diversion from the already underfunded malaria programs (89,105) Tailoring these interventions to local contexts and scaling up through community-based approaches, including health education and promotion, environmental cleanup, and leveraging the role of gender dynamics in household-level vector control, is also critical for ensuring their sustainability and success (51,93,104,106) Additionally, strengthening these strategies through multisectoral collaboration across agriculture, environment, urban planning, education and public health, would foster a more holistic and coordinated approach to arboviral threats nationwide (106).

To reinforce Kenya’s response to arboviral outbreaks, developing and refining targeted contingency plans is essential. Kenya has made some progress in this regard with the RVFV contingency plan, which has been periodically updated to focus on high-risk areas such as Isiolo and Wajir (107). These plans should be further adapted by pre-positioning resources in these regions, training healthcare workers in rapid response protocols, and ensuring the IDSR system is fully equipped to handle the nuances of arboviral outbreaks (108,109). Kenya can also capitalize our findings to contribute to a national dedicated arbovirus dashboard— per GAI’s priority actions—as a comprehensive tool for real-time monitoring, optimal resource allocation and improved coordination during outbreaks(7,9)Further, multicountry collaborations through regional networks like the East African Community and African Union, alongside global partners through the GAI, could provide Kenya with additional resources, expertise and data to strengthen its capacity to manage local and cross-border threats, while also supporting continent-level early warning systems (7,93).

Our findings may however be limited by substantial heterogeneity, as sources of heterogeneity, such as varying methodologies, population characteristics, and confirmation cut-offs were not reported in many studies and could not be assessed. Additionally, our county-level data, based on sampled areas, may not fully represent local ecological and climatic diversity, necessitating caution in extrapolating findings to entire counties. Ascertainment bias may also be present, as included studies predominantly focused in identified “hotspot” areas, potentially overrepresenting them. Finally, reliance on ELISA without PRNT or RT-PCR confirmation across most studies raises concerns on results accuracy and reliability. Nonetheless, our study reveals an extensive longstanding history of arboviral circulation in Kenya, with future risks projected to increase due to ecological, environmental, and socioeconomic factors. For effective mitigation, strengthening surveillance, enhancing integrated vector management, and improving outbreak response and prevention efforts are key. By addressing these key areas through targeted interventions and regional cooperation, Kenya can better position itself to effectively manage the growing risk of arboviral diseases.

## Supporting information

S1 PRISMA Checklist

S2 Literature Search Syntax

S3 JBI Critical Appraisal Checklist

S4 Data Extraction Form

S5 Quality Appraisal Outputs

S6 Study characteristics

## Data Availability

All relevant data has been included in the manuscript and supplementary material.

## Contributors

L.J.K. conceived the study, conducted the systematic search, and was the first reviewer for the entire study. R.N., E.K., H.A. and J.L. contributed to screening, data extraction and quality appraisal. L.J.K. and J.L. conducted the statistical analysis, and L.J.K. wrote the first draft of the manuscript with input from J.L. and H.A., B.B., J.A. and D.M. reviewed the manuscript and gave input on the statistical analysis and interpretation of results. All authors contributed to drafting and revising the manuscript.

## Data sharing

All relevant data has been included in the manuscript and supplementary material.

## Acknowledgements

The authors express their gratitude for the funding by DTRA HDTRA11910031. D.M.M was supported by the ILRI/BMZ One Health Research, Education, Outreach and Awareness Centre (OHRECA) program. The funders had no role in study design, data collection and analysis, decision to publish, or preparation of the manuscript.

## Supplementary materials

