## Supplementary material for "Arboviruses in Kenya: A Systematic Review and Meta-analysis of Prevalence": S2 Literature Search Syntax

**Search Date: 15/03/2024**

### PubMed

| Query | Syntax | Hits |
|---|---|---|
| #1 | Epidemiology OR Prevalence OR Seroprevalence OR Distribution OR Risk factors OR Seroepidemiology OR Surveillance | 5,807,793 |
| #2 | Vector-borne Virus* OR Arthropod-borne OR Arthropod-borne Virus* OR Mosquito-borne Virus* OR Aedes OR Culex OR Anopheles OR Tick-borne Virus* OR Arbovir* OR Yellow fever OR YF OR Rift Valley Fever Virus OR RVFV OR Dengue OR DENV OR Chikungunya OR CHIKV OR Zika OR West Nile Virus OR WNV OR Crimean-Congo Hemorrhagic Fever OR CCHF OR Mayaro Virus* OR O'nyong' nyong OR O’nyong’ nyong OR O’nyong nyong OR ONNV OR Thogoto OR Nairobi Sheep Disease OR Tick-borne Encephalitis Virus* OR Bunyavir* OR Alphavir* OR Flavivir* OR Phlebovir* OR Nairovir* OR Togovir* OR Orthomyxovir* OR Orthobunyavir* | 143,297 |
| #3 | Kenya | 41,358 |
| **#4** | **#1 AND #2 AND #3** | **928** |

### Global Health

| Query | Syntax | Hits |
|---|---|---|
| #1 | Epidemiology OR Prevalence OR Seroprevalence OR Distribution OR Risk factors OR Seroepidemiology OR Surveillance | 1,092,679 |
| #2 | Vector-borne Virus* OR Arthropod-borne OR Arthropod-borne Virus* OR Mosquito-borne Virus* OR Aedes OR Culex OR Anopheles OR Tick-borne Virus* OR Arbovir* OR Yellow fever OR YF OR Rift Valley Fever Virus OR RVFV OR Dengue OR DENV OR Chikungunya OR CHIKV OR Zika OR West Nile Virus OR WNV OR Crimean-Congo Hemorrhagic Fever OR CCHF OR Mayaro Virus* OR O'nyong' nyong OR O’nyong’ nyong OR O’nyong nyong OR ONNV OR Thogoto OR Nairobi Sheep Disease OR Tick-borne Encephalitis Virus* OR Bunyavir* OR Alphavir* OR Flavivir* OR Phlebovir* OR Nairovir* OR Togovir* OR Orthomyxovir* OR Orthobunyavir* | 184,984 |
| #3 | Kenya | 23,142 |
| **#4** | **#1 AND #2 AND #3** | **851** |

### Web of Science

| Query | Syntax | Hits |
|---|---|---|
| #1 | Epidemiology OR Prevalence OR Seroprevalence OR Distribution OR Risk factors OR Seroepidemiology OR Surveillance | 6,092,854 |
| #2 | Vector-borne OR Arthropod-borne Virus* OR Mosquito-borne Virus* OR Aedes OR Culex OR Anopheles OR Tick-borne Virus* OR Arbovir* OR Yellow fever OR Rift Valley Fever Virus OR Dengue OR Chikungunya OR Zika OR West Nile Virus OR Crimean-Congo Hemorrhagic Fever OR Mayaro Virus* OR O'nyong' nyong OR O’nyong nyong OR Thogoto OR Nairobi Sheep Disease OR Tick-borne Encephalitis Virus* OR Bunyavir* OR Alphavir* OR Flavivir* OR Phlebovir* OR Nairovir* OR Togovir* OR Orthomyxovir* OR Orthobunyavir* | 128,595 |
| #3 | Kenya | 91,533 |
| **#4** | **#1 AND #2 AND #3** | **1,306** |
