## Supplementary material for "Arboviruses in Kenya: A Systematic Review and Meta-analysis of Prevalence": S3 JBI Critical Appraisal Checklist

### CHECKLIST FOR PREVALENCE STUDIES

Critical Appraisal tools for use in JBI Systematic Reviews

jbi.global

CRICOS Provider Number 00123M

### INTRODUCTION

JBI is an international research organisation based in the Faculty of Health and Medical Sciences at the University of Adelaide, South Australia. JBI develops and delivers unique evidence-based information, software, education and training designed to improve healthcare practice and health outcomes. With over 70 Collaborating Entities, servicing over 90 countries, JBI is a recognised global leader in evidence-based healthcare.

#### JBI Systematic Reviews

The core of evidence synthesis is the systematic review of literature of a particular intervention, condition or issue. The systematic review is essentially an analysis of the available literature (that is, evidence) and a judgment of the effectiveness or otherwise of a practice, involving a series of complex steps. JBI takes a particular view on what counts as evidence and the methods utilised to synthesise those different types of evidence. In line with this broader view of evidence, JBI has developed theories, methodologies and rigorous processes for the critical appraisal and synthesis of these diverse forms of evidence in order to aid in clinical decision-making in healthcare. There now exists JBI guidance for conducting reviews of effectiveness research, qualitative research, prevalence/incidence, etiology/risk, economic evaluations, text/opinion, diagnostic test accuracy, mixed-methods, umbrella reviews and scoping reviews. Further information regarding JBI systematic reviews can be found in the JBI Evidence Synthesis Manual.

#### JBI Critical Appraisal Tools

All systematic reviews incorporate a process of critique or appraisal of the research evidence. The purpose of this appraisal is to assess the methodological quality of a study and to determine the extent to which a study has addressed the possibility of bias in its design, conduct and analysis. All papers selected for inclusion in the systematic review (that is – those that meet the inclusion criteria described in the protocol) need to be subjected to rigorous appraisal by two critical appraisers. The results of this appraisal can then be used to inform synthesis and interpretation of the results of the study. JBI Critical appraisal tools have been developed by the JBI and collaborators and approved by the JBI Scientific Committee following extensive peer review. Although designed for use in systematic reviews, JBI critical appraisal tools can also be used when creating Critically Appraised Topics (CAT), in journal clubs and as an educational tool.

© JBI, 2020. All rights reserved. JBI grants use of these tools for research purposes only. All other enquiries should be sent to.

### JBI CRITICAL APPRAISAL CHECKLIST FOR STUDIES REPORTING PREVALENCE DATA

Reviewer_______________________________________ Date_______________________________

Author________________________________________Year_________ Record Number_________

| | | Yes | No | Unclear | Not applicable |
|---|---|---|---|---|---|
| 1. | Was the sample frame appropriate to address the target population? | ☐ | ☐ | ☐ | ☐ |
| 2. | Were study participants sampled in an appropriate way? | ☐ | ☐ | ☐ | ☐ |
| 3. | Was the sample size adequate? | ☐ | ☐ | ☐ | ☐ |
| 4. | Were the study subjects and the setting described in detail? | ☐ | ☐ | ☐ | ☐ |
| 5. | Was the data analysis conducted with sufficient coverage of the identified sample? | ☐ | ☐ | ☐ | ☐ |
| 6. | Were valid methods used for the identification of the condition? | ☐ | ☐ | ☐ | ☐ |
| 7. | Was the condition measured in a standard, reliable way for all participants? | ☐ | ☐ | ☐ | ☐ |
| 8. | Was there appropriate statistical analysis? | ☐ | ☐ | ☐ | ☐ |
| 9. | Was the response rate adequate, and if not, was the low response rate managed appropriately? | ☐ | ☐ | ☐ | ☐ |

Overall appraisal: Include ☐ Exclude ☐ Seek further info ☐

Comments (Including reason for exclusion)

________________________________________________________________________________

________________________________________________________________________________

© JBI, 2020. All rights reserved. JBI grants use of these tools for research purposes only. All other enquiries should be sent to.

### JBI CRITICAL APPRAISAL CHECKLIST FOR STUDIES REPORTING PREVALENCE DATA

*How to cite: Munn Z, Moola S, Lisy K, Riitano D, Tufanaru C. Methodological guidance for systematic reviews of observational epidemiological studies reporting prevalence and incidence data. Int J Evid Based Healthc. 2015;13(3):147–153.*

Answers: Yes, No, Unclear or Not/Applicable

1. **Was the sample frame appropriate to address the target population?**

   This question relies upon knowledge of the broader characteristics of the population of interest and the geographical area. If the study is of women with breast cancer, knowledge of at least the characteristics, demographics and medical history is needed. The term "target population" should not be taken to infer every individual from everywhere or with similar disease or exposure characteristics. Instead, give consideration to specific population characteristics in the study, including age range, gender, morbidities, medications, and other potentially influential factors. For example, a sample frame may not be appropriate to address the target population if a certain group has been used (such as those working for one organisation, or one profession) and the results then inferred to the target population (i.e. working adults). A sample frame may be appropriate when it includes almost all the members of the target population (i.e. a census, or a complete list of participants or complete registry data).

2. **Were study participants recruited in an appropriate way?**

   Studies may report random sampling from a population, and the methods section should report how sampling was performed. Random probabilistic sampling from a defined subset of the population (sample frame) should be employed in most cases, however, random probabilistic sampling is not needed when everyone in the sampling frame will be included/ analysed. For example, reporting on all the data from a good census is appropriate as a good census will identify everybody. When using cluster sampling, such as a random sample of villages within a region, the methods need to be clearly stated as the precision of the final prevalence estimate incorporates the clustering effect. Convenience samples, such as a street survey or interviewing lots of people at a public gatherings are not considered to provide a representative sample of the base population.

© JBI, 2020. All rights reserved. JBI grants use of these tools for research purposes only. All other enquiries should be sent to.

3. **Was the sample size adequate?**

The larger the sample, the narrower will be the confidence interval around the prevalence estimate, making the results more precise. An adequate sample size is important to ensure good precision of the final estimate. Ideally we are looking for evidence that the authors conducted a sample size calculation to determine an adequate sample size. This will estimate how many subjects are needed to produce a reliable estimate of the measure(s) of interest. For conditions with a low prevalence, a larger sample size is needed. Also consider sample sizes for subgroup (or characteristics) analyses, and whether these are appropriate. Sometimes, the study will be large enough (as in large national surveys) whereby a sample size calculation is not required. In these cases, sample size can be considered adequate.

When there is no sample size calculation and it is not a large national survey, the reviewers may consider conducting their own sample size analysis using the following formula: (Naing et al. 2006, Daniel 1999)

n= Z2P(1-P)

d2

Where:

n= sample size

Z = Z statistic for a level of confidence

P = Expected prevalence or proportion (in proportion of one; if 20%, P = 0.2)

d = precision (in proportion of one; if 5%, d=0.05)

**Ref:**

Naing L, Winn T, Rusli BN. Practical issues in calculating the sample size for prevalence studies Archives of Orofacial Sciences. 2006;1:9-14.

Daniel WW. Biostatistics: A Foundation for Analysis in the Health Sciences.

Edition. 7th ed. New York: John Wiley & Sons. 1999.

© JBI, 2020. All rights reserved. JBI grants use of these tools for research purposes only. All other enquiries should be sent to.

4. **Were the study subjects and setting described in detail?**

   Certain diseases or conditions vary in prevalence across different geographic regions and populations (e.g. Women vs. Men, sociodemographic variables between countries). The study sample should be described in sufficient detail so that other researchers can determine if it is comparable to the population of interest to them.

5. **Was data analysis conducted with sufficient coverage of the identified sample?**

   Coverage bias can occur when not all subgroups of the identified sample respond at the same rate. For instance, you may have a very high response rate overall for your study, but the response rate for a certain subgroup (i.e. older adults) may be quite low.

6. **Were valid methods used for the identification of the condition?**

   Here we are looking for measurement or classification bias. Many health problems are not easily diagnosed or defined and some measures may not be capable of including or excluding appropriate levels or stages of the health problem. If the outcomes were assessed based on existing definitions or diagnostic criteria, then the answer to this question is likely to be yes. If the outcomes were assessed using observer reported, or self-reported scales, the risk of over- or under-reporting is increased, and objectivity is compromised. Importantly, determine if the measurement tools used were validated instruments as this has a significant impact on outcome assessment validity.

7. **Was the condition measured in a standard, reliable way for all participants?**

   Considerable judgment is required to determine the presence of some health outcomes. Having established the validity of the outcome measurement instrument (see item 6 of this scale), it is important to establish how the measurement was conducted. Were those involved in collecting data trained or educated in the use of the instrument/s? If there was more than one data collector, were they similar in terms of level of education, clinical or research experience, or level of responsibility in the piece of research being appraised? When there was more than one observer or collector, was there comparison of results from across the observers? Was the condition measured in the same way for all participants?

© JBI, 2020. All rights reserved. JBI grants use of these tools for research purposes only. All other enquiries should be sent to.

8. **Was there appropriate statistical analysis?**

   Importantly, the numerator and denominator should be clearly reported, and percentages should be given with confidence intervals. The methods section should be detailed enough for reviewers to identify the analytical technique used and how specific variables were measured. Additionally, it is also important to assess the appropriateness of the analytical strategy in terms of the assumptions associated with the approach as differing methods of analysis are based on differing assumptions about the data and how it will respond.

9. **Was the response rate adequate, and if not, was the low response rate managed appropriately?**

   A large number of dropouts, refusals or "not founds" amongst selected subjects may diminish a study's validity, as can a low response rates for survey studies. The authors should clearly discuss the response rate and any reasons for non-response and compare persons in the study to those not in the study, particularly with regards to their socio-demographic characteristics. If reasons for non-response appear to be unrelated to the outcome measured and the characteristics of non-responders are comparable to those who do respond in the study (addressed in question 5, coverage bias), the researchers may be able to justify a more modest response rate.

© JBI, 2020. All rights reserved. JBI grants use of these tools for research purposes only. All other enquiries should be sent to.
